# Sex differences in the association between age-related decline in blood pressure and decline in cognition: A prospective cohort study

**DOI:** 10.1101/2025.01.08.25320209

**Authors:** Nicholas W. Baumgartner, Ana W. Capuano, Lisa L. Barnes, David A. Bennett, Zoe Arvanitakis

## Abstract

**Background:** Both high and declining blood pressure (BP) are associated with cognitive decline risk in older adults. In late-life, women have higher rates of hypertension, experience faster cognitive decline, and represent two-thirds of individuals with Alzheimer’s disease dementia. However, sex differences in the association between BP decline and cognitive decline are unknown.

**Methods:** Data were analyzed from 4719 older adults without known baseline dementia (mean age = 76.7 [SD = 7.7] years; 74% women) enrolled in one of five US-based prospective community-based cohort studies, followed annually for up to 31 years (mean = 8.7 [SD = 5.7] years). A 19-test cognitive battery, yielding composite global and five domain-specific scores, and BP were assessed annually. Bivariate mixed-effects models simultaneously estimated change in BP and cognition, for the total group and by sex.

**Findings:** Systolic BP, diastolic BP, and cognition all declined over time (ps <0.01). Bivariate mixed-effect models revealed a sex difference in the correlation of decline in systolic BP and decline in global cognition (women: r = 0.26, 95%CI: 0.17 - 0.37; men: r = 0.01, 95%CI: -0.13 - 0.11), such that women exhibited a stronger correlation than men. Decline in systolic BP was related to decline in global and all five cognitive domains in women but none in men, with another sex difference identified in the working memory domain. An increase of diastolic BP was related to decline in working memory in men, and no other associations with diastolic BP were significant for either sex.

**Interpretation:** Systolic BP decline in late-life is related to decline in global and domain-specific cognition in women but not men, with sex differences in global cognition and the working memory domain. These findings suggest that in older women, declining systolic BP – a routinely-used clinical measure – may be an important marker of concurrent cognitive decline.

## Introduction

High blood pressure (BP) and cognitive impairment are common at older ages^1^. Numerous cross-sectional and longitudinal studies have indicated both midlife (ages 40–64 years) and late-life hypertension (ages 65+) are associated with increased risk of cognitive decline^2^, although some evidence suggests that low BP in late-life may also increase the risk of cognitive impairment^2^. Hypertension is a key modifiable risk factor for cerebrovascular pathology^3^, which itself is a major cause of cognitive decline and dementia in old age^4^. However, decline in systolic BP (SBP) has also been associated with an increased risk of dementia^5^ and worse performance on the Mini-Mental State Exam (MMSE)^6,7^. Likewise, our prior study of 1288 older deceased participants indicated that higher levels of SBP and diastolic BP (DBP), as well as a faster decline in SBP, were each associated with a greater number of infarcts in the postmortem brain^8^.

To date, sex differences in the association between BP and cognition remain unclear. Men have higher rates of hypertension in early to middle adulthood, but women have higher rates after age 65 years^9^ and women comprise nearly two-thirds of all persons affected by Alzheimer’s disease^10^. These patterns suggest that BP and cognitive changes disproportionately affect women in later life. However, the relationship between declining BP and declining cognitive function over years – and whether these relationships differ by sex – is unknown. Understanding these relationships could enhance precision medicine by informing interventions that target BP to mitigate risks associated with cognitive decline in older age. Investigating sex differences may also help reduce health disparities, improve patient outcomes, and guide better clinical decision-making.

In this study, we investigated the relationship between long-term changes in BP and cognitive decline in late-life, with a focus on identifying sex differences. Utilizing data from five longitudinal community-based cohorts of more than 4700 older adults, we assessed how changes in SBP and DBP over time were related to cognitive decline on a global measure and across five domains. The findings aim to deepen our understanding of sex-specific associations linking BP and cognition, with implications for targeted intervention strategies.

## Methods

### Cohorts and Study Design

Participants were drawn from five ongoing longitudinal, prospective, community-based cohort studies of aging, the Religious Orders Study (ROS)^11^, the Rush Memory and Aging Project (MAP)^12^, the Minority Aging Research Study (MARS)^13^, the Rush Alzheimer’s Disease Center African American Clinical Core (AA core)^14^, and the Latino Core (LATC)^15^. These studies share a similar design and standardized data collection, enabling the combination of harmonized data from the five cohorts. Each study was approved by a Rush institutional Review Board of Rush University Medical Center, and all participants provided written informed consent before enrollment.

The ROS began in 1994 by enrolling Catholic nuns, priests, and brothers from about 40 convents and monasteries across the United States. MAP, initiated in 1997, consists of older community members recruited from retirement and subsidized housing facilities across the Chicagoland area. Both MARS (since 2004) and the AA Core (since 2008) consist of older Black adults while LATC (since 2015) enrolls older Latino/Hispanic adults, all recruited from churches, retirement homes, senior buildings, and social clubs in Chicago neighborhoods and surrounding suburbs. Follow-up rates across the five cohorts are high, ranging from 85% to 90%^11–13^.

All studies recruit participants without known dementia at baseline. Annual evaluations are harmonized to include medical history, physical examination with vital signs, height/weight measurements, neuropsychological testing, and neurological evaluation. Annual follow-up visits are identical to baseline protocols. Medications (including antihypertensive medications) and over-the-counter agents (e.g., vitamins, supplements) taken within 2 weeks preceding the annual evaluation are visually inspected and documented, including name and dosage. Demographics including age (from date of birth), sex (male/female), years of education, history of hypertension, age at menopause, and race/ethnicity are self-reported.

### Participants

At time of analysis, eligible participants had no missing data at baseline for the main covariates of interest, and at least two measurements – baseline and at least one other – of composite global cognition, SBP, and DBP. Of the 5394 participants who completed their baseline visit, 673 were ineligible due to recent enrollment (e.g., only baseline data available; n = 465), incomplete baseline data (n = 79), or missing data (n = 129;122 of which had only one BP measurement). To ensure comparability across sexes, participants were restricted to those aged 50-100 years old at baseline (n = 2 removed). The final analytic sample comprised 4719 participants.

### Procedures

BP was measured at baseline and annually by a trained researcher using an automated sphygmomanometer. Three readings **–** two seated and one standing **–** were taken roughly one minute apart at each visit. Standing BP was not significantly different from sitting; therefore, the three measurements were averaged to calculate mean SBP and DBP. For statistical analyses, SBP and DBP values were standardized into z-scores using the baseline mean and standard deviation of the entire cohort. Clinical evaluations documented demographics (self-reported sex and racioethnic status), medical history including self-reported history of hypertension and age at menopause, medications including antihypertensives (verified by visual inspection), and other measures such as body mass index (BMI) calculated from the height and weight.

Cognitive performance was assessed annually using a comprehensive battery of 19 different neuropsychological tests, as detailed previously^16^. The tests evaluated a broad spectrum of cognitive abilities organized into a composite measure of global cognition and five specific cognitive domains. Higher scores indicated better cognitive performance. Briefly, three tests of working memory (Digit Span forward and backward; Digit Ordering), three of semantic memory (Boston Naming; Verbal Fluency; Reading Test), seven of episodic memory (Word List Memory, Recall and Recognition; immediate and delayed recall of the East Boston Story; immediate and delayed recall of Story A of the Wechsler Memory Scale-Revised), four of perceptual speed (Symbol Digit Modalities Test; Number Comparison; two indices from a modified version of the Stroop Test), and two tests of visuospatial abilities (Line Orientation; Progressive Matrices) were administered. Raw scores were standardized into z-scores using the baseline mean and standard deviation of the entire cohort. Composite scores for each domain were calculated by averaging the z-scores for all tests within that domain. The global cognitive function composite score was derived by averaging z-scores across all 19 tests.

### Statistical Analysis

A descriptive summary of baseline variables was conducted to characterize the sample. All models were adjusted for age (centered at 76.7 years), education (centered at 15.8 years), and antihypertensive medication use (1 = yes, 0 = no) as well as interactions of these variables with time. Models not stratified by sex were also adjusted for sex and its interaction with time. Linear mixed-effects models were used to examine changes over time, with SBP, DBP, global cognition, and five cognitive domains as outcomes. Next, to determine whether change in BP was related to change in cognition, simultaneous bivariate mixed-effects regression analyses with both BP (SBP and DBP modeled separately) and cognition (global cognition and each domain separately) modeled simultaneously as outcomes were used. The models were then repeated stratified by sex. Fully adjusted bivariate mixed-effect models estimated simultaneously the levels and rates of change in BP and cognition, while the association (i.e. covariance) of these levels and rates of change in both outcomes were characterized by a joint distribution of the four random effects (i.e., the covariance between the two intercepts and two slopes). From the derived covariance-covariance G-matrix (see **Supplemental Table S1**), the correlation between level and the correlation between rate of change were calculated for all outcomes. Sex differences were determined in two steps. First, confidence intervals for the correlations between rates of change ascertained using the bivariate mixed effect model were calculated through Fisher transformations. Second, significant differences in correlation were confirmed using bootstrapping analysis (50 runs, mean 30 hours run total), a more time-consuming but conservative approach. Bootstrap confidence intervals are reported. Sensitivity analyses repeated the fully adjusted simultaneous bivariate mixed-effects regression analyses in the total sample and stratified by sex, adding BMI, race/ethnicity, or age at menopause in separate models, along with their interactions with time. All analyses were conducted using SAS/STAT software, version 9.4 of the SAS system for Linux (SAS Institute, Cary, NC), with α set at 0.05.

## Results

A descriptive summary of baseline variables in the total group and stratified by sex is reported in **Table 1**. Adjusted linear mixed-effect models (**Table 2**) revealed a significant effect of time on SBP, DBP, and all cognitive measures, indicating these measures declined over time for the whole sample and for both sexes separately (ps <0.01). Men experienced a faster decline in DBP than women (p <0.01; **Figure 1**), as indicated by a significant interaction between sex and time in fully adjusted models. However, no sex differences were observed in the rate of decline in SBP or decline in cognition.

**Figure 1.**
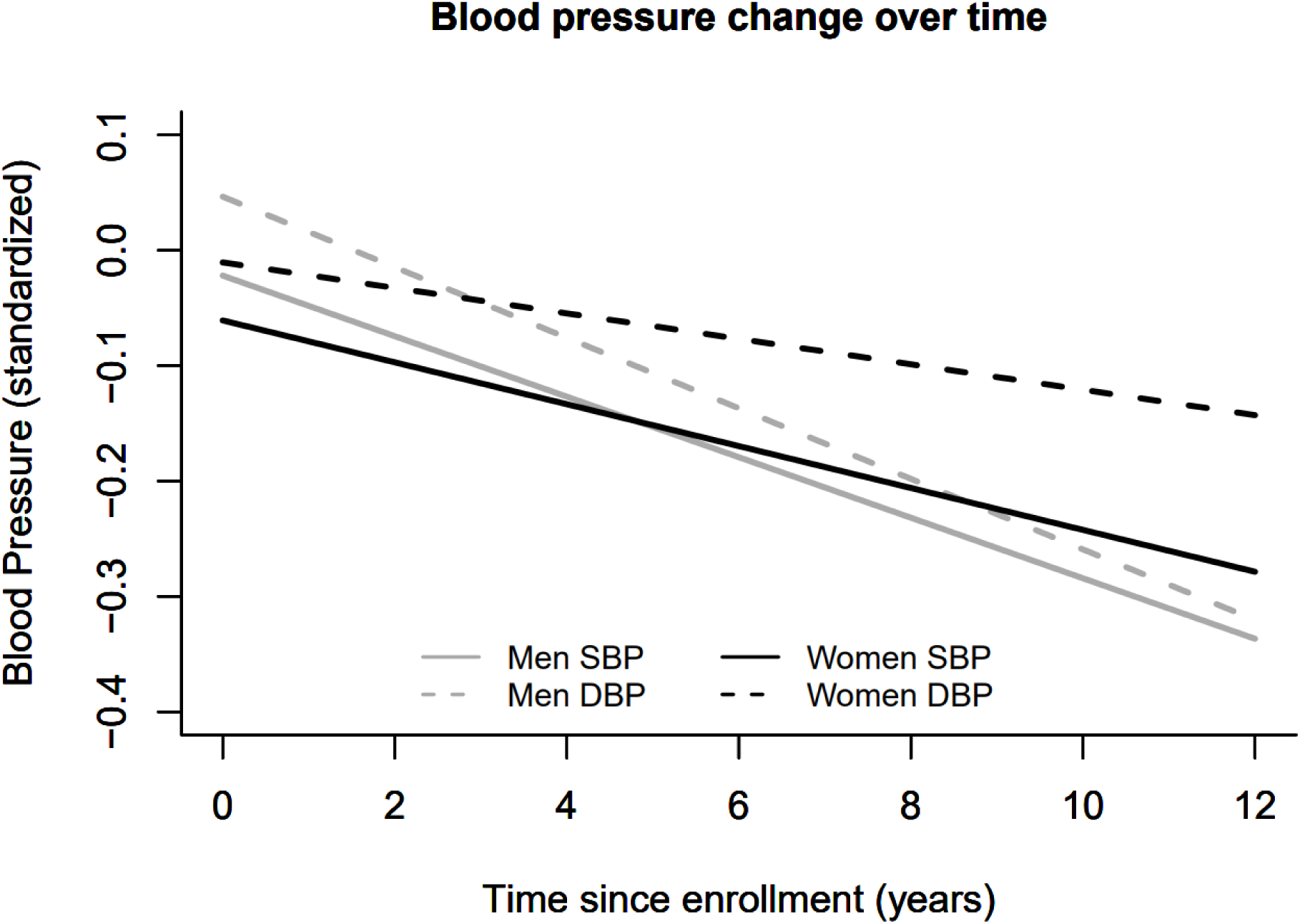
Fully adjusted linear mixed effect models show longitudinal trajectories of SBP (solid line) and DBP (dashed line) separately for women (black) and men (grey). Blood pressure is presented in standardized units (z-score using the baseline mean and standard deviation of the entire cohort). Both SBP and DBP decreased significantly over time (p <0.01). While SBP decline rates did not differ by sex, men exhibited a faster DBP decline rate than women (p <0.01). Covariates included age, sex, education, antihypertensive use, and their interactions with time.

**Table 1.** Analytic Sample Characteristics.

|  | **Total**<br>**(n = 4719)** | **Women**<br>**(n = 3502)** | **Men**<br>**(n = 1217)** |
| --- | --- | --- | --- |
| Demographic |  |  |  |
| Age (years) | 76.7 ± 7.7 | 76.7 ± 7.8 | 76.8 ± 7.5 |
| Race/Ethnicity (n, %) |  |  |  |
| White, non-Latino | 3057, 64.8% | 2201, 62.9% | 856, 70.3% |
| Black, non-Latino | 1243, 26.3% | 976, 27.9% | 267, 22.0% |
| Latino | 387, 8.2% | 305, 8.7% | 82, 6.7% |
| Other | 32, 0.7% | 20, 0.6% | 12, 1.0% |
| Education (years) | 15.8 ± 3.9 | 15.5 ± 3.8 | 16.5 ± 4.3 |
| Annual follow up visits (years) | 8.7 ± 5.7 | 8.8 ± 5.6 | 8.2 ± 5.8 |
| Clinical |  |  |  |
| Systolic blood pressure (mmHg) | 134.9 ± 19.0 | 135.2 ± 19.3 | 134.0 ± 18.3 |
| Diastolic blood pressure (mmHg) | 76.2 ± 11.5 | 76.4 ± 11.6 | 75.8 ± 11.4 |
| Systolic blood pressure slope (mmHg/year) | -0.02 ± 0.04 | -0.02 ± 0.04 | -0.02 ± 0.03 |
| Diastolic blood pressure slope (mmHg/year) | -0.02 ± 0.03 | -0.02 ± 0.03 | -0.02 ± 0.04 |
| History of hypertension (n, %) | 2644, 56.0% | 2034, 58.1% | 610, 50.1% |
| Antihypertensive medication use (n, %) | 3142, 66.6% | 2322, 66.3% | 820, 67.4% |
| Body Mass Index (kg/m<sup>2</sup>) | 28.2 ± 5.5 | 28.3 ± 5.9 | 27.93 ± 4.3 |
| Age at menopause (years) | 46.8 ± 7.0 | 46.8 ± 7.0 | n/a |
| Cognition |  |  |  |
| Global cognitive function score | 0.03 ± 0.62 | 0.06 ± 0.60 | -0.04 ± 0.66 |
| Working memory | 0.03 ± 0.78 | 0.03 ± 0.78 | 0.02 ± 0.79 |
| Semantic memory | 0.04 ± 0.81 | 0.06 ± 0.81 | -0.02 ± 0.81 |
| Episodic memory | 0.04 ± 0.74 | 0.09 ± 0.72 | -0.12 ± 0.79 |
| Perceptual speed | 0.05 ± 0.83 | 0.11 ± 0.81 | -0.10 ± 0.86 |
| Visuospatial ability | 0.03 ± 0.82 | -0.04 ± 0.79 | 0.23 ± 0.85 |

Values are mean ± standard deviation unless otherwise noted; all variables represent value at first baseline visit unless otherwise specified.

**Table 2.**
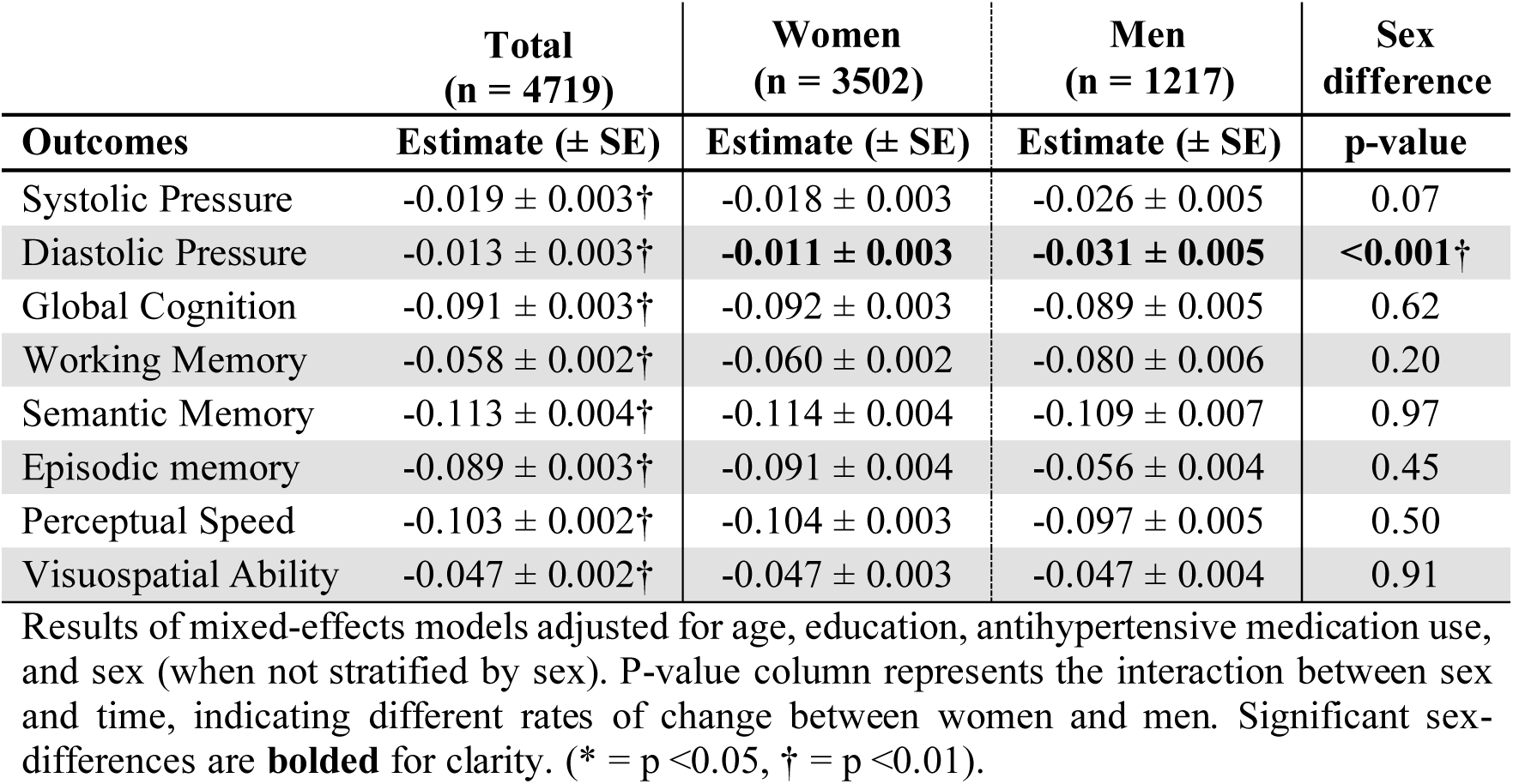
Estimates of Change in Blood Pressure and Change in Cognition.

Both levels (intercept terms) and slopes (time terms) of SBP were associated with global cognition in fully adjusted simultaneous bivariate mixed-effects regression analyses modeling BP and global cognition across the full sample (**Supplemental Table S2**). Specifically, participants with higher levels of SBP had lower global cognition performance, while those with a faster decline in SBP had faster declines in global cognition. Although higher DBP levels were associated with lower global cognition, declines in DBP were not related to changes in cognition.

Models were repeated stratified by sex to assess associations between BP and cognition separately for women and men, reported in **Table 3**. In women, level of SBP was not related to level of global cognition, however, decline of SBP was related to decline in global cognition. In men, higher levels of SBP were associated with lower global cognition scores but change in SBP was not. Women exhibited a stronger correlation between decline in SBP and decline in global cognition (r = 0.26, 95%CI: 0.17 - 0.37) than men (r = 0.01, 95%CI: -0.13 - 0.11) in correlations obtained from adjusted simultaneous linear mixed effect models with bootstrap confidence intervals calculated within sex, as shown in **Figure 2**. For DBP, higher levels were associated with lower levels of global cognition in both sexes but change in DBP were not. No significant sex differences were observed for DBP.

**Figure 2.**
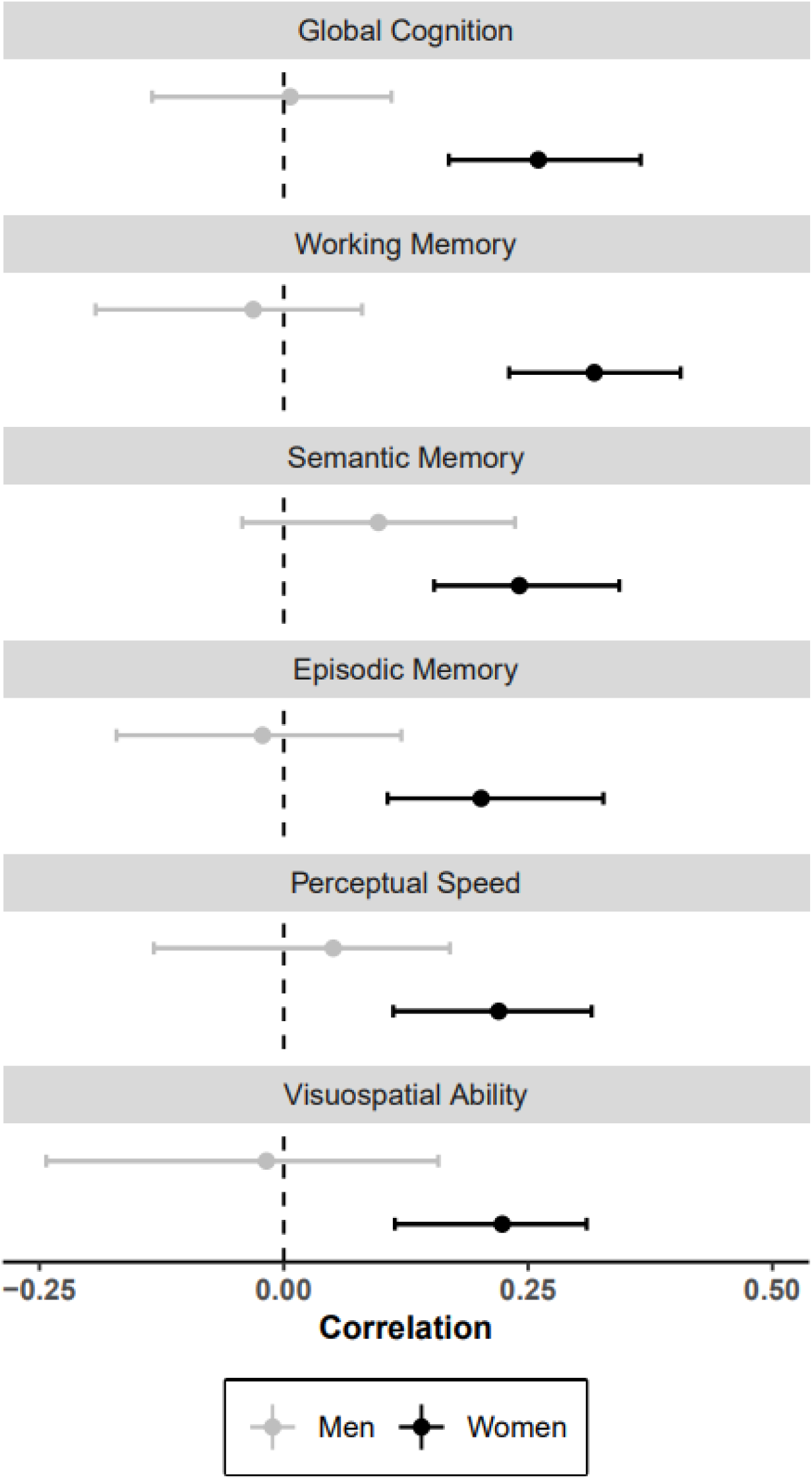
Correlation between change in SBP and change in cognition calculated from the simultaneous fully adjusted bivariate mixed-effects regression models for global cognition and the five separate domains. Women (black) showed significant correlations between decline in SBP and decline in global and all five domains, while correlations for men (grey) were not statistically different from zero. Bootstrapped 95% confidence intervals confirmed sex differences, such that women had stronger correlations between decline in SBP and decline in global cognition, as well as with decline in working memory, compared to men.

**Table 3.**
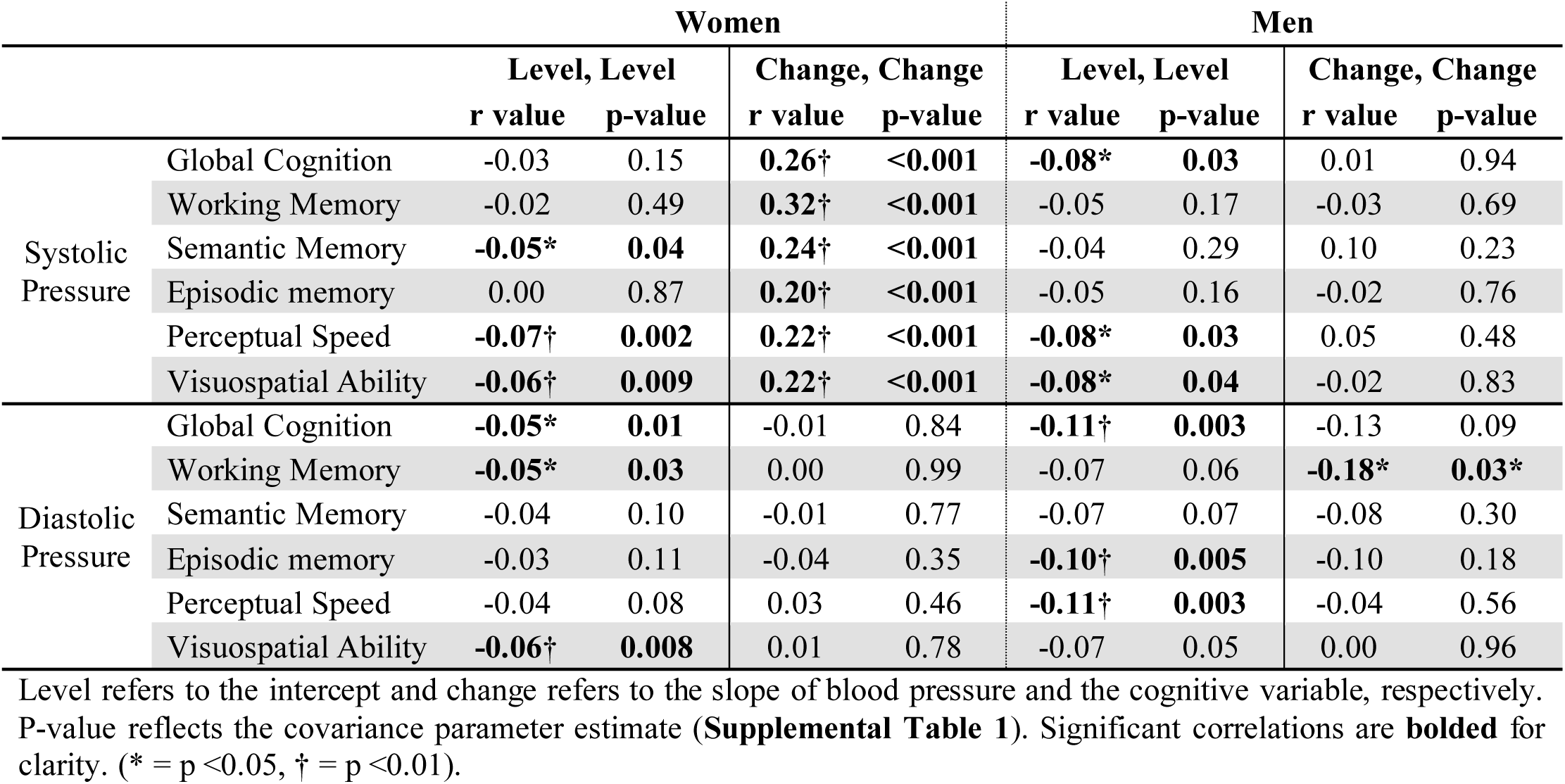
Correlations of Level and Change of Blood Pressure with Level and Change of Cognition and Five Cognitive Domains by Sex.

Higher levels of SBP were inversely related to levels of semantic memory, perceptual speed, and visuospatial abilities, but not working memory or episodic memory performance in simultaneous adjusted bivariate mixed-effects regression models (**Supplemental Table S2**) modeling BP and cognition across five separate domains. Faster decline in SBP was related to faster declines across all five domains. Higher levels of DBP were associated with lower levels of performance across all five domains, but changes in DBP were not related to changes in any cognitive domains.

Analyses repeated stratified by sex (**Table 3**) showed that in women, higher level of SBP was associated with lower level of semantic memory, perceptual speed and visuospatial abilities, but not working memory or episodic memory. Women with faster decline in SBP had faster declines in all five cognitive domains. Conversely, men with higher levels of SBP tended to have worse perceptual speed and visuospatial ability, but changes in SPB were not related to changes in any cognitive domains. Women exhibited a stronger correlation between decline in SBP and decline in working memory (r = 0.32, 95%CI: 0.23 - 0.41) compared to men (r = -0.03, 95%CI: -0.19 - 0.08), as shown in **Figure 2**. No other sex differences were observed. For DBP, in women higher levels of DBP were associated with lower levels of working memory and visuospatial ability, but not other domains. Change in DBP was not related to any domains for women. For men, higher levels of DBP were associated with lower episodic memory and perceptual speed. Increasing DBP was related to faster decline in working memory for men, but no other relationships with other cognitive domains were observed for men. No significant sex differences were identified for DBP.

Past studies have shown that individuals with both obesity and hypertension tend to have worse cognitive function compared to those with just hypertension^17^ and the impact of higher SBP on decline in cognition is greater in Blacks compared to Whites^18^. Similarly, menopause-related hormonal changes have been linked to increased risk of hypertension and cognitive decline^19^. Sensitivity analyses were conducted modeling change in SBP and change in global cognition. The addition of BMI, race/ethnicity, or age at menopause did not change the results of any models, suggesting they do not explain the effects.

## Discussion

In this community-based longitudinal study of more than 4700 older adults examined annually for an average of 9 years, we identified significant associations between simultaneous decline in SBP and decline in global cognitive function. Pre-specified sex-stratified analyses revealed that a faster decline in SBP was associated with a faster decline in global cognition among women but not men, with a significant sex difference observed. Secondary analyses showed that SBP decline was related to declines across five cognitive domains in the total cohort; however, when stratified by sex, the relationship only existed in women. A significant sex difference was observed in the working memory domain, such that the relationship was stronger for women than for men. Conversely a faster increase in DBP was associated with a faster decline in working memory in men but not women. As the medical field moves towards precision medicine, our findings suggest declining SBP – a measurement routinely and repeatedly monitored in clinics worldwide – may be an important correlate of concurrent cognitive decline in women.

Past research, primarily using cross-sectional designs, has linked hypertension to worse cognitive function in older adults^2^, impacting various cognitive domains including immediate and delayed memory, episodic memory, executive function, processing speed, and visuospatial abilities^2,18^ among others. Results of our cross-sectional analyses largely support prior research, finding that higher baseline SBP and DBP were associated with worse global cognitive function. Given the higher prevalence of hypertension^9^ and dementia^10^ in older women, researchers have increasingly been exploring sex differences in BP and cognition. Previous works have reported higher BP levels are associated with worse cognitive function, but across different domains for both women and men separately^20^. Our results are in keeping with the literature, as higher levels of SBP and DBP were associated with worse cognitive function for both sexes separately, albeit not uniformly across all cognitive domains. Collectively, these results reinforce the association between higher levels of BP and lower levels of cognitive function in older adults.

Few studies have assessed how change in BP relates to concurrent change in cognitive function, with most longitudinal research focusing on the relationship between higher BP at a single time point with worse cognitive outcomes years later. For instance, higher baseline SBP or DBP have been shown to predict worse processing speed, visuospatial ability, immediate memory, and executive function 8-13 years later^21–23^. On the other hand, a limited number of studies using a less precise measure of cognition have found that *low* baseline BP was associated with lower MMSE scores at follow up^22,24^. These data point to the complex relation of level of BP to level of cognition years later, however, few studies have evaluated change in cognition. Longitudinal studies with repeated BP and cognitive assessments have found that sustained hypertension (SBP >150 mmHg) was associated with greater cognitive decline compared to normotensive individuals^25^. Conversely, it has been reported that low baseline BP is associated with faster annual decline in MMSE scores^26^, while decline in SBP measured at baseline and a single follow up was associated with decline in MMSE scores over the same period^6,7^. Notably, studies reporting an association between decline in BP and decline in cognition have solely used cognitive screening tools (i.e. MMSE) rather than comprehensive assessments of global and domain-specific cognition. This study expands our understanding by reporting for the first time that the decline in SBP is related to concurrent decline in global cognition and across five cognitive domains using composite measures assessed annually. These findings suggest that declining SBP in older adults may signal broader cognitive decline across multiple domains, advocating for closer monitoring of routine BP trends as an integral part of cognitive health assessments.

Older women experience higher rates of hypertension^9^ and dementia^10^, despite this, the sex-specific relationship between changes in BP and changes in cognition remains largely unexplored. Our sex-stratified analysis indicated that decline in SBP was related to declines in global cognition and all five cognitive domains for women, whereas no relationships were observed in men. Significant sex differences were identified, with women showing stronger correlations between decline in SBP and declines in global cognition, as well as in the domain of working memory, compared to men who showed no associations. Although few, prior studies generally suggest that *higher* SBP – not declining SBP – in women is related to worsening performance across several cognitive domains at follow-up, with no relationships in men^27,28^. Notably, one study reported a decline in SBP from baseline was associated with a higher probability of decreased MMSE scores three years later, an association that persisted only for women in sex-stratified analyses^6^. However, the results may be limited by the study’s small sample of men (n = 223)^6^, the MMSE’s reduced sensitivity, and a short follow-up period. Nonetheless, our study extends and reinforces and extends this association by linking decline in SBP and declines across multiple cognitive domains in women and confirming sex differences for global cognition as well as within the working memory domain. In contrast, our study identified that faster increase in DBP was related to a faster decline in working memory in men but not women. While previous research on DBP has yielded mixed results^2^, our study did not detect sex differences in these associations. To our knowledge, no prior studies have reported sex differences in the relationship between DBP and working memory, though one study in men only reported lower DBP at age 50 predicted better working memory performance 20 years later^29^, in keeping with our results. Together, our findings suggest that monitoring BP changes, particularly SBP in women, could serve as a practical marker identifying concurrent cognitive decline. Since BP is routinely measured at medical visits – while cognitive testing is more time consuming and occurs only after concerns arise – tracking SBP trends in older women may support earlier identification of cognitive decline. Proactive monitoring and interventions based on BP trends could improve long-term outcomes for those most at risk.

Several plausible factors may explain the observed relationship between change in BP and change in cognition. Medical (i.e. BMI^17^) and socio-demographic (i.e. race/ethnicity^18^) factors have been shown to influence this association, but adjusting for these did not alter our findings. Age related variation in the association between BP and cognition^2^ was unlikely to influence the results, as the sample included a wide range of older persons and models accounted for age as a covariate. Chronic hypertension, a risk factor for cerebral damage and arterial stiffening, impairs cerebral blood flow, which can contribute to cognitive decline^30^. Therefore, elevated SBP in late-life may offer some protection against cognitive decline by maintaining cerebral blood flow in the face of arteriosclerosis-induced resistance^31^, whereas declining BP may indicate impaired compensatory mechanisms needed to maintain cerebral perfusion.

Several factors may contribute to the sex differences observed. Sex differences may arise from menopause-related hormonal changes, which increase risks of hypertension and cognitive decline^19^, though adjusting for age at menopause did not change the results. While prior studies reported small male sample sizes limited their ability to detect sex-differences^6^, our large male sample (n = 1,217) ensured sufficient statistical power. Lastly, disparities in hypertension detection and treatment – such as women being less likely to receive antihypertensive medication^32^, seek urgent medical care^33^, or discuss cardiovascular risk factors with their physicians^33^ – may result in poorer BP management in women over the lifespan, amplifying the impact of BP changes on cognition.

This study had several limitations. First, like many aging cohorts, our sample primarily consisted of White women, potentially limiting generalizability. The observational design also introduces possible biases inherent in the sample, such as survival bias, particularly given that high SBP is the leading cause of death globally^34^. However, this concern is partially mitigated by the consistency of our associations regardless of baseline BP or level throughout the study. Despite these limitations, this study possesses notable strengths. We leveraged a large, longitudinal cohort with over 31 years of annual data collection, enabling detailed assessment of changes over time. The comprehensive design included annual BP measurements and a comprehensive battery of 19 neuropsychological tests across five cognitive domains, allowing robust analyses of cognitive and BP trajectories over multiple time points. The use of a composite cognitive score reduced ceiling and floor effects compared to screening tools like the MMSE. Additionally, the inclusion of five distinct cognitive domains offered valuable insights into the domains most affected by changing BP. Finally, simultaneous mixed-effects regression analyses enabled us to examine both the levels and changes in BP and cognition simultaneously, leveraging multidimensional longitudinal data to draw more robust conclusions.

## Contributors

**NWB:** Conceptualization, Writing - Original Draft, and Writing - Review & Editing. **AWC:** Conceptualization, Data curation, Formal analysis, Methodology, Supervision, Validation, Visualization, Writing - Review & Editing. **LLB:** Resources, and Writing - Review & Editing. **DAB:** Resources, and Writing - Review & Editing. **ZA:** Conceptualization, Funding acquisition, Methodology, Project administration, Resources, Supervision, and Writing - Review & Editing.

## Data availability

Raw data are available upon request from qualified investigators applying through the Rush Alzheimer’s Disease Center (RADC) Research Resource Sharing Hub (radc.rush.edu/home.htm).

## Acknowledgments

This work was supported by the National Institutes of Health grant numbers RF1AG074549, R01AG017917, R01AG15819, R01AG022018, P30AG010161, and P30AG072975. The authors would like to thank the entire staff at the Rush Alzheimer’s Disease Center.

---

**Table S1.** Covariance Parameter Estimates of Change in Blood Pressure with Change in Cognition and Five Cognitive Domains by Sex.

|  |  | Women |  |  |  | Men |  |  |  |
| --- | --- | --- | --- | --- | --- | --- | --- | --- | --- |
| Bivariate Outcomes | | Estimate (± SE) | Z Value | Lower | Upper | Estimate (± SE) | Z Value | Lower | Upper |
| Systolic Pressure | Global Cognition | **0.00133 ± 0.00021** | **6.22<sup>†</sup>** | **0.00091** | **0.00175** | 0.00003 ± 0.00037 | 0.09 | -0.00069 | 0.00075 |
| | Working Memory | **0.00109 ± 0.00016** | **6.88<sup>†</sup>** | **0.00078** | **0.00140** | -0.00010 ± 0.00025 | -0.40 | -0.00058 | 0.00039 |
| | Semantic Memory | **0.00157 ± 0.00028** | **5.60<sup>†</sup>** | **0.00102** | **0.00212** | 0.00061 ± 0.00050 | 1.21 | -0.00038 | 0.00159 |
| | Episodic memory | **0.00119 ± 0.00024** | **4.89<sup>†</sup>** | **0.00071** | **0.00167** | -0.00012 ± 0.00040 | -0.30 | -0.00090 | 0.00066 |
| | Perceptual Speed | **0.00086 ± 0.00017** | **4.96<sup>†</sup>** | **0.00052** | **0.00120** | 0.00019 ± 0.00027 | 0.71 | -0.00033 | 0.00071 |
| | Visuospatial Ability | **0.00071 ± 0.00016** | **4.48<sup>†</sup>** | **0.00040** | **0.00103** | -0.00005 ± 0.00025 | -0.22 | -0.00054 | 0.00043 |
| Diastolic Pressure | Global Cognition | -0.00004 ± 0.00020 | -0.20 | -0.00044 | 0.00036 | -0.00063 ± 0.00038 | -1.66 | -0.00137 | 0.00011 |
| | Working Memory | 0.00000 ± 0.00015 | 0.01 | -0.00029 | 0.00029 | **-0.00058 ± 0.00026** | **-2.24\*** | **-0.00109** | **-0.00007** |
| | Semantic Memory | -0.00008 ± 0.00027 | -0.29 | -0.00060 | 0.00045 | -0.00053 ± 0.00052 | -1.04 | -0.00154 | 0.00047 |
| | Episodic memory | -0.00022 ± 0.00023 | -0.94 | -0.00067 | 0.00024 | -0.00055 ± 0.00041 | -1.35 | -0.00136 | 0.00025 |
| | Perceptual Speed | 0.00012 ± 0.00017 | 0.75 | -0.00020 | 0.00045 | -0.00016 ± 0.00028 | -0.58 | -0.00070 | 0.00038 |
| | Visuospatial Ability | 0.00004 ± 0.00015 | 0.28 | -0.00025 | 0.00034 | -0.00001 ± 0.00026 | -0.04 | -0.00052 | 0.00050 |

Results of bivariate mixed-effects models jointly modeling both change in blood pressure and cognition over time, adjusted for age, education, and antihypertensive medication use. Lower and upper bounds of 95%CI from bivariate mixed-effects models are presented for each variable. Significant results are **bolded** for clarity. (\* = $p < 0.05$ , <sup>†</sup> = $p < 0.01$ ).

**Table S2.** Correlations of Level and Change of Blood Pressure with Level and Change of Cognition and Five Cognitive Domains.

|  |  | Level, Level |  | Change, Change |  |
| --- | --- | --- | --- | --- | --- |
|  |  | r value | p-value | r value | p-value |
| Systolic Pressure | Global Cognition | **-0.04\*** | **0.04** | **0.20†** | **<0.001** |
|  | Working Memory | -0.02 | 0.31 | **0.24†** | **<0.001** |
|  | Semantic Memory | **-0.04\*** | **0.04** | **0.21†** | **<0.001** |
|  | Episodic memory | -0.01 | 0.61 | **0.15†** | **<0.001** |
|  | Perceptual Speed | **-0.07†** | **<0.001** | **0.18†** | **<0.001** |
|  | Visuospatial Ability | **-0.06†** | **0.002** | **0.16†** | **<0.001** |
| Diastolic Pressure | Global Cognition | **-0.06†** | **0.007** | -0.03 | 0.35 |
|  | Working Memory | **-0.05†** | **0.008** | -0.04 | 0.28 |
|  | Semantic Memory | **-0.04\*** | **0.03** | -0.03 | 0.45 |
|  | Episodic memory | **-0.05\*** | **0.01** | -0.05 | 0.15 |
|  | Perceptual Speed | **-0.05†** | **0.004** | -0.01 | 0.79 |
|  | Visuospatial Ability | **-0.06†** | **0.002** | 0.01 | 0.84 |

Level indicates the intercept and change indicates the slope of blood pressure and the cognitive variable. P-value reflects the covariance parameter estimate (data not shown). Significant correlations are **bolded** for clarity (\* = $p < 0.05$ , † = $p < 0.01$ )

